# Differential Cytokine Signatures of SARS-CoV-2 and Influenza Infection Highlight Key Differences in Pathobiology

**DOI:** 10.1101/2021.01.29.21250317

**Authors:** Andrew H. Karaba, Weiqiang Zhou, Leon L. Hsieh, Alexis Figueroa, Guido Massaccesi, Richard E. Rothman, Katherine Z.J. Fenstermacher, Lauren Sauer, Kathryn Shaw-Saliba, Paul W. Blair, Sherry Leung, Russell Wesson, Nada Alachkar, Ramy El-Diwany, Hongkai Ji, Andrea L. Cox

## Abstract

**Background:** Several inflammatory cytokines are upregulated in severe COVID-19. We compared cytokines in COVID-19 versus influenza in order to define differentiating features of the inflammatory response to these pathogens and their association with severe disease. Because elevated body mass index (BMI) is a known risk factor for severe COVID-19, we examined the relationship of BMI to cytokines associated with severe disease.

**Methods:** Thirty-seven cytokines and chemokines were measured in plasma from 145 patients with COVID-19, 57 patients with influenza, and 30 healthy controls. Controlling for BMI, age, and sex, differences in cytokines between groups were determined by linear regression and random forest prediction was utilized to determine the cytokines most important in distinguishing severe COVID-19 and influenza. Mediation analysis was utilized to identify cytokines that mediate the effect of BMI on disease severity.

**Results:** IL-18, IL-1β, IL-6, and TNF-α were significantly increased in COVID-19 versus influenza patients while GM-CSF, IFN-γ, IFN-λ1, IL-10, IL-15, and MCP-2 were significantly elevated in the influenza group. In subgroup analysis based on disease severity, IL-18, IL-6, and TNF-α were elevated in severe COVID-19, but not severe influenza. Random forest analysis identified high IL-6 and low IFN-λ1 levels as the most distinct between severe COVID-19 and severe influenza. Finally, IL-1RA was identified as a potential mediator of the effects of BMI on COVID-19 severity.

**Conclusions:** These findings point to activation of fundamentally different innate immune pathways in SARS-CoV-2 and influenza infection, and emphasize drivers of severe COVID-19 to focus both mechanistic and therapeutic investigations.

**Summary:** Severe COVID-19 is marked by dysregulated inflammation and is associated with elevated BMI. By comparing cytokines and chemokines in patients with either COVID-19 or influenza, we identified distinct inflammatory pathways and a cytokine mediator of the effect of BMI.

## INTRODUCTION

Severe Acute Respiratory Syndrome Coronavirus 2 (SARS-CoV-2) has infected more than 90 million people and led to more than 2 million deaths worldwide in 2020 *[1]*. COVID-19, the disease caused by SARS-CoV-2, spans mild disease with few symptoms to multiorgan failure and death *[2,3]*. One hallmark of severe disease is immune dysregulation characterized by elevated proinflammatory markers and inflammatory cytokines[4–9] including interleukin (IL)-6, IL-10, IP-10, IL-1RA, and MCP-1 [8,10–15]. These findings led to trials of targeted anti-inflammatory therapies such as IL-6 and IL-1 antagonists[16–20]. Retrospective analyses of these agents and randomized controlled trials demonstrate modest benefit [21–26]. However, studies have called into question the uniqueness of the inflammatory cytokine profile of COVID-19 by highlighting similarities to sepsis or acute respiratory distress syndrome (ARDS) due to other causes [27–29].

Influenza is another viral respiratory infection capable of causing severe pneumonia and pandemics [30]. The case fatality rate for influenza is lower than that of COVID-19, but many of the same cytokines upregulated in COVID-19 are also increased in severe influenza infection [15,31,32]. Thus, it is unclear what is unique to cytokine upregulation in SARS-CoV-2 infection that leads to more severe disease than with other viruses [33]. Several clinical factors correlate with severe COVID-19, including advanced age and elevated BMI [34,35]. Obese patients are at increased risk for hospitalization and death, particularly at younger ages [36]. Obesity leads to chronic inflammation, and elevated BMI is specifically associated with increases in IL-10, IL-6, TNF-α, and IL-1RA [37–39]. Yet, few studies examining these cytokines in COVID-19 have incorporated this in their analyses.

As SARS-CoV-2 infections continue through the 2020-2021 influenza season, people will be exposed to both viruses. To determine how the cytokines produced during these two infections differ and to understand the increased pathogenicity of SARS-CoV-2, we measured thirty-seven cytokines and chemokines in patients hospitalized with either influenza or COVID-19 and compared the levels of these inflammatory mediators based on disease severity. We also performed mediation analysis to identify cytokines that mediate the effect of BMI on severity of disease. We found that severe COVID-19 induces a macrophage proinflammatory cytokine profile, while severe influenza leads to interferon pathway induction. We also found that IL-1RA, produced by adipocytes as well as immune cells, is a mediator potentially explaining the underlying relationship between obesity and severe COVID-19. These findings highlight that disparate immune pathways are activated in these potentially life-threatening respiratory viral infections.

## METHODS

### Study Participants and Samples

All studies were approved by the Johns Hopkins (JH) Institutional Review Board. Hospitalized patients diagnosed with COVID-19 by positive SARS-CoV-2 RNA testing in the Johns Hopkins Healthcare System were enrolled in a prospective consented protocol to investigate research questions specific to the clinical course of COVID-19 (JH IRB 00245545). Demographic information, clinical laboratory test results, ICD-10 coded diagnoses (comorbidities), BMI, and other clinical parameters were linked to data for all COVID-19 patients in the study. Participants were categorized by maximum COVID-19 disease severity score based on the WHO severity scale [40]. Those with a score of 5 or lower were categorized as having mild/moderate disease and those with a 6 or higher were considered severe. Blood was obtained as close to admission as feasible and centrifuged at 400 x g for 5 min to separate cells from plasma in BSL2+ laboratory conditions. Plasma was frozen at −80°C until thawed for cytokine measurement as described below.

The healthy control (HC) plasma samples were obtained from HIV/HCV-antibody seronegative subjects enrolled before 2020 in the Baltimore Before and After Acute Study of Hepatitis (BBAASH) study (JH IRB NA_00046368), an ongoing prospective, community-recruited, observational cohort study of people who inject drugs as previously described [41].

Plasma samples from hospitalized patients infected with influenza between 2017 and 2019 were obtained as previously described for comparison to hospitalized COVID-19 patients in this study [41,42], under an IRB approved protocol (JH IRB 00091667). Patients hospitalized with influenza requiring no more than nasal cannula and those that required higher levels of oxygen support were classified as having mild/moderate disease or severe disease, respectively, which approximates the WHO COVID-19 severity score.

### Cytokine measurement

Plasma was thawed and cytokines and chemokines (IFN-α2a, IFN-β, IL-18, IL-1RA, IL-23, IFN-λ1, IL-2Ra, MCP-2, GM-CSF, IL-23p40, IL-15, IL-16, IL-17A, IL-1α, IL-5, IL-7, TNF-β, VEGF, Eotaxin, Eotaxin-3, IP-10, MCP-1, MCP-4, MDC, MIP-1α, MIP-1β, TARC, IFN-γ, IL-10, IL-12p70, IL-13, IL-1β, IL-2, IL-4, IL-6, IL-8, TNF-α) were measured using a custom multiplex kit from Meso Scale Discovery (MSD, Rockville, MD) according to the manufacture’s protocol and data were acquired on a MESO QuickPlex SQ 120. Each sample was measured on first thaw and in duplicate. If an analyte signal was below background, it was set to 0 and if detectable, but below the manufacturer’s internal lower limit of quantification, to the lower limit of detection.

### Statistical Analysis

Data were analyzed using the statistical computing software R version 3.6.3 [43]. The cytokine/chemokine (i.e., analyte) signals were first log_2_ transformed after adding a pseudocount of one. To compare the analytes between patient groups a linear regression analysis, which is equivalent to a two-tailed t-test after adjusting for covariates, was applied. For instance, a linear regression model was fitted for each analyte to test the difference between every pair of patient groups (e.g., COVID-19 vs. influenza) after adjusting for covariates (age, gender, and BMI). P-values of the coefficient for the patient group from the model were obtained and converted to false discovery rates (FDR) using the Benjamini-Hochberg (BH) procedure[44]. An FDR of 0.25 was considered significant. Pearson’s correlation between each pair of analytes was calculated based on the data from both the COVID-19 and influenza subjects or based on data from each disease separately. Random forest and mediation analysis were performed as described in the supplementary methods [45–47].

## RESULTS

### Patient Characteristics

A total of 145 SARS-CoV-2 infected participants, 57 influenza infected participants, and 30 HCs were studied. We separated the influenza and COVID-19 groups based on final infection outcome into mild/moderate and severe, as described in Materials and Methods. Thirteen out of 57 (23%) participants in the influenza cohort and 89 out of 145 (61%) COVID-19 patients had severe disease (Table 1). The influenza and COVID-19 cohorts were not significantly different in gender, non-white race, or BMI. The influenza cohort was younger than the COVID-19 cohort on average (mean age 48.2 versus 56.6 years).

**Table 1.**
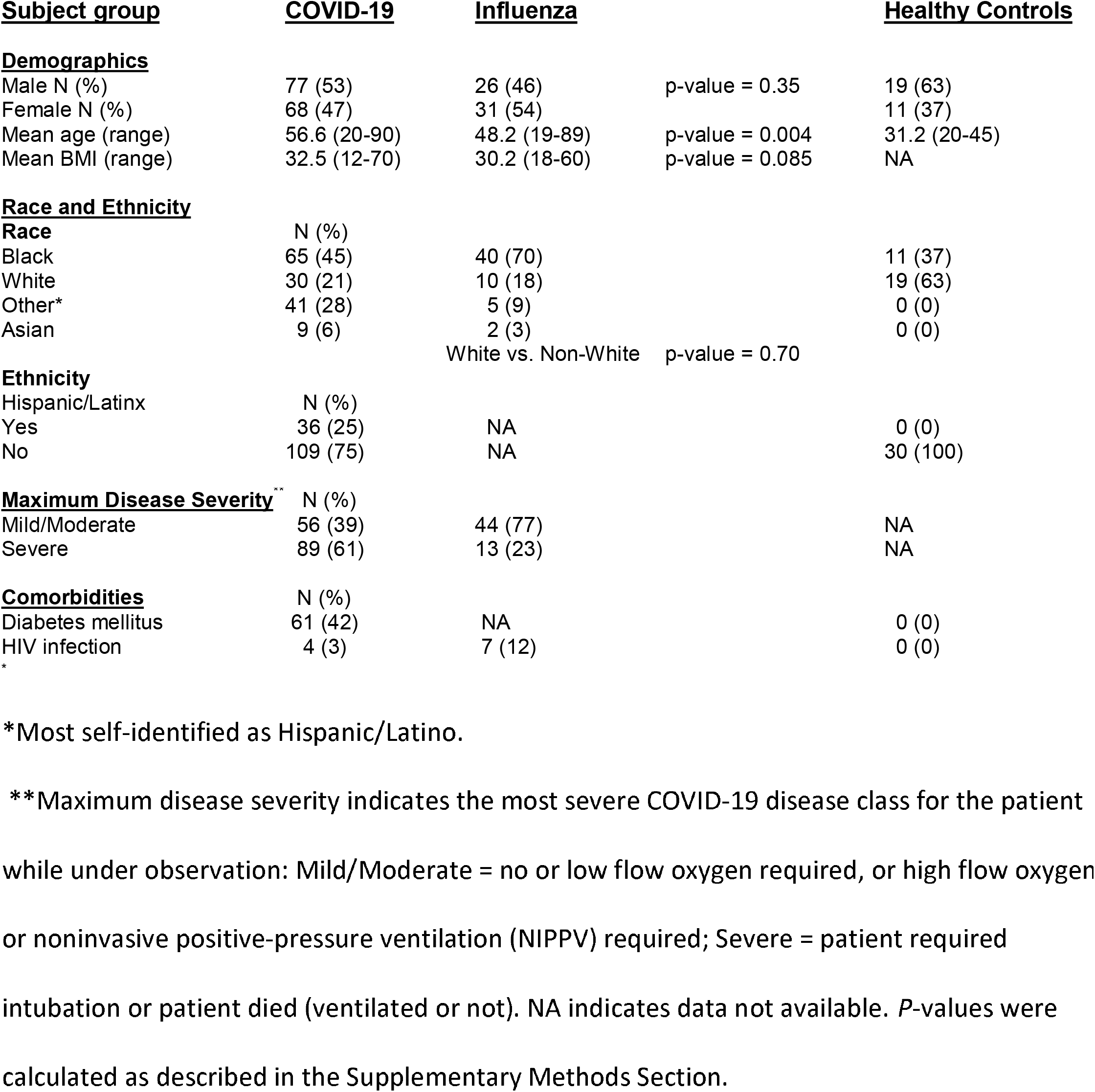
Characteristics of study subjects.

### Cytokine Elevations in COVID-19 and Influenza Compared to Healthy Controls

To determine which cytokines and chemokines are upregulated in influenza and COVID-19, we measured cytokines/chemokines in HCs and compared them to those with influenza and COVID-19 at time of study enrollment. The analytes chosen in our custom panel were based on prior publications in SARS-CoV-1, SARS-CoV-2, and influenza demonstrating their potential as important markers of disease severity [8,15,48–50]. We found that GM-CSF, IFN-β, IFN-γ, IFN-λ1, IL-10, IL-15, IL-18, IL-1RA, IL-6, IL-8, IP-10, MCP-1, MCP-2, and TNF-α were significantly increased in both influenza and COVID-19 compared to HCs (**Figure 1**). In contrast, IL-13, eotaxin-3, MDC, and TARC were significantly decreased in COVID-19 and influenza compared to HCs. Six analytes were not significantly elevated above HCs in either virus group: IL-1α, IL-23, IL-5, MCP-4, MIP-1α, and MIP-1β (**Supplemental Figure 1**). Additionally, IL-1β, IL-2, IL-23p40, IL-2Ra, IL-7, and VEGF were elevated in COVID-19 exclusively, not influenza, compared to HCs. Conversely, IFN-α2a, IL-16, and IL-17A were significantly elevated compared to the HCs solely in the influenza cohort, with no difference between COVID-19 participants and HCs.

**Figure 1.**
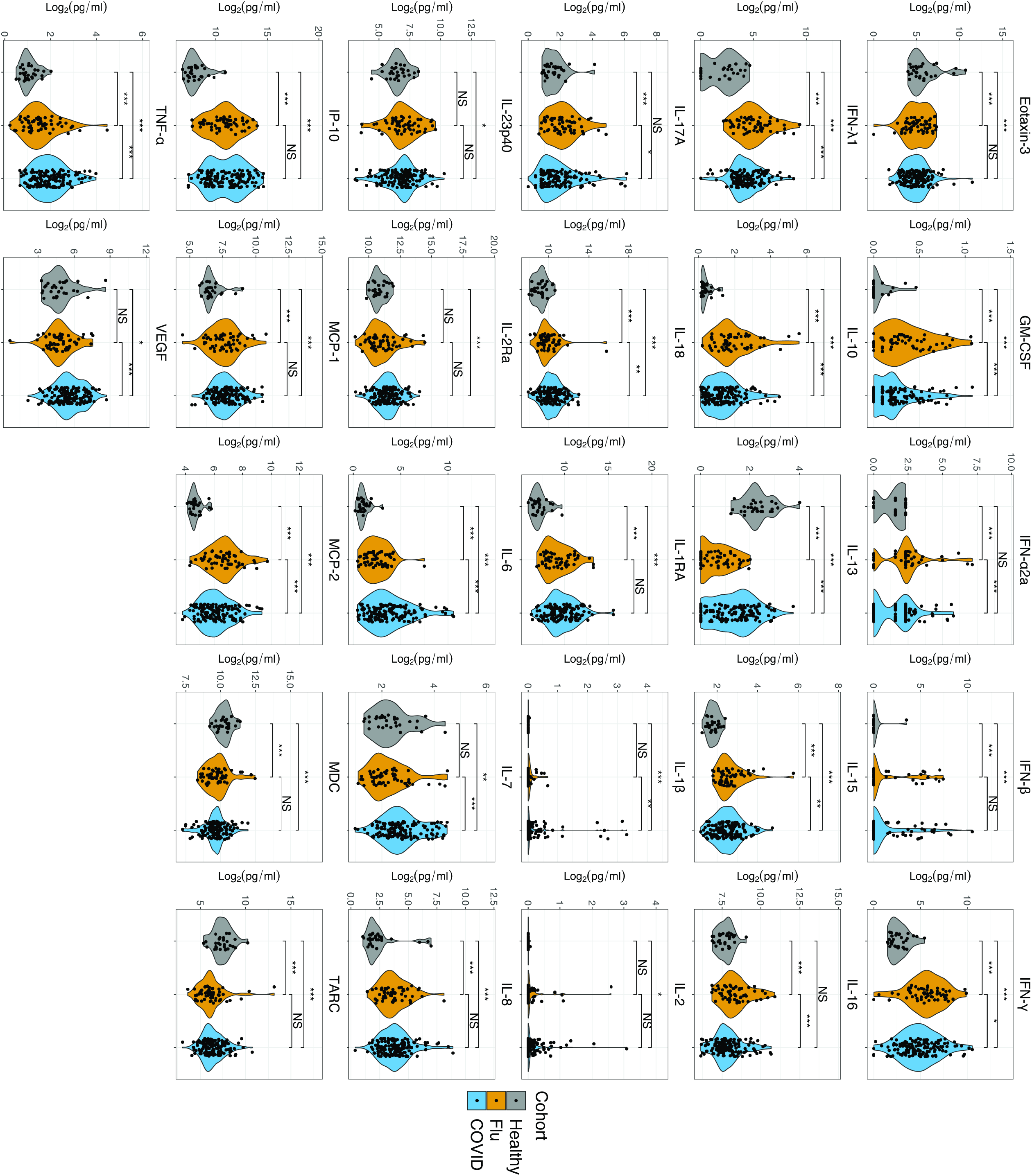
Cytokines and Chemokines in Influenza and COVID-19 Compared to Healthy Controls. Differences between the COVID-19 cohort (blue) or influenza cohort (orange) and healthy controls (grey) were determined by two-tailed t-test after adjusting for sex and age. Differences between the COVID-19 cohort and influenza cohort for each analyte were determined by two-tailed t-test after adjusting for sex, age, and BMI. FDR was obtained using the Benjamini-Hochberg procedure. Statistical significance is indicated by NS, *, **, or *** above the brackets indicating FDR >0.25, <0.25, <0.1, or <0.05 respectively.

### Differences in Cytokines and Chemokines Between Influenza and COVID-19 Reveal a Proinflammatory Macrophage Signature in COVID-19

We then compared each cytokine or chemokine in the COVID-19 cohort to the influenza cohort at time of enrollment. IL-18, IL-1β, IL-6, IL-7, TNF-α, IL-2Ra, and VEGF were higher in COVID-19, while GM-CSF, IFN-γ, IFN-λ1, IL-10, IL-15, and MCP-2 were higher in influenza (**Figure 1**). When adjusted for BMI, IL-2Ra was no longer significantly different between the two infections (**Supplementary Table 1 and 2**). The cytokines most elevated in the COVID-19 group are produced primarily by macrophages and also characterize macrophage activation syndrome (MAS) [51,52]. Elevated levels of both IL-18 and IL-1β suggest prominent inflammasome activation in COVID-19 compared to influenza [53]. Macrophages are major sources of inflammasome cytokines in other viral infections [54–56]. Notably, IFN-λ1 was nearly 2-fold higher in influenza compared to COVID-19, consistent with a study that identified limited induction of IFNs-λ1, −α2a, and −β *in vitro* by SARS-CoV-2 [49], and a small study in humans demonstrating a paucity of interferon production in COVID-19 versus influenza [57]. Though IL-10 was implicated in COVID-19 pathogenesis [15,58,59], IL-10 levels were actually higher in moderate influenza compared to COVID-19.

Plotting the correlation of each analyte with every other analyte in correlation matrices by disease revealed that many of the cytokines/chemokines elevated in influenza relative to COVID-19 strongly correlate, including IL-10, IFN-λ1, MCP-2, and IFN-γ. Similarly, those increased in COVID-19 relative to influenza positively correlate including IL-18, TNF-α, and IL-6 (**Figure 2A and 2B**). This suggests that two distinct inflammatory pathways are activated in these respiratory viral infections. Next, we generated a heatmap grouped by disease subgroup (**Figure 2C)**. Several patterns emerged, including increases in IL-6, TNF-α, IL-18, and IL-1RA in severe COVID-19 compared to both mild/moderate COVID-19 and influenza. In contrast, IFN-α2a and IFN-λ1 were higher in the influenza cohort compared to COVID-19.

**Figure 2.**
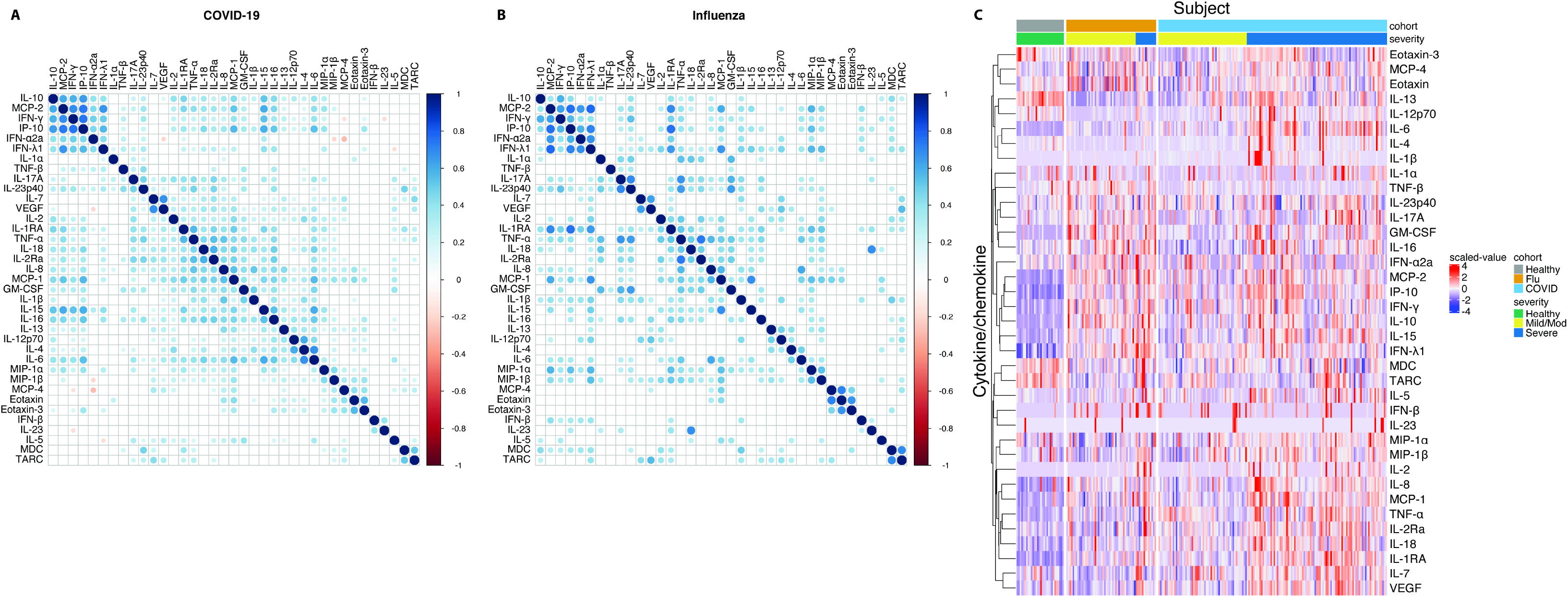
Cytokine and Chemokine Correlation Matrix and Heatmap of Cytokines and Chemokines Grouped by Disease Subgroup. A and B. Correlations between each analyte and every other analyte were determined by Pearson’s correlation coefficient of each pair of cytokine/chemokine across all COVID-19 (**A**) and influenza (**B**) patients. The heat bar on the right side of the figure indicates both the directionality of the correlation (blue represents a positive correlation and red represents a negative correlation) and the strength (darker values indicate a correlation coefficient closer to 1 or −1). **<u>C</u>**. Each row represents an individual analyte and each column represents an individual in the study. The healthy controls are on the far left of the figure and marked by grey and green bars at the top of the figure. Participants with influenza are in the middle and marked with an orange bar at the top of the figure. The COVID-19 cohort is on the right half of the figure and marked by a light blue bar at the top of the figure. A yellow bar indicates those with mild disease (influenza or COVID-19) and a dark blue bar indicates severe disease (influenza or COVID-19). The scaled values were computed by standardizing the signals of each analyte to have zero mean and standard deviation of one across all participants and are represented by the key on the right of the figure where darker red represents values with a greater fold change above the average and darker blue represents values with a greater decrease below the average.

When we compared these analytes statistically by severity subgroups, we found minimal overlap in the cytokines/chemokines that distinguished severe from mild/moderate influenza and those that distinguished severe from mild/moderate COVID-19 (**Figure 3A and 3B**). With and without adjustment for BMI, IL-1RA, IL-1β, IL-2, IL-7, MCP-2, and VEGF were elevated in both severe diseases compared to their mild/moderate counterpart. MCP-1 was also significantly elevated in both groups when adjusting for BMI. With and without adjustment for BMI, only IFN-β and IFN-λ1 were elevated in severe influenza, but not in severe COVID-19 (**Figure 3 and Supplementary Table 1 and 2**), consistent with low interferon responses in COVID-19. Cytokines elevated in severe COVID-19, but not severe influenza, include GM-CSF, IL-10, IL-15, IL-18, IL-2Ra, IL-6, IL-8, IP-10, and TNF-α (**Figure 3A and 3B**). Of the cytokines elevated in severe COVID-19, but not severe influenza, only IL-18, IL-6, and TNF-α were also elevated in the whole COVID-19 cohort compared to the whole influenza cohort (**Figures 1 and 3B**). These three cytokines are highly associated with proinflammatory macrophages[60].

**Figure 3.**
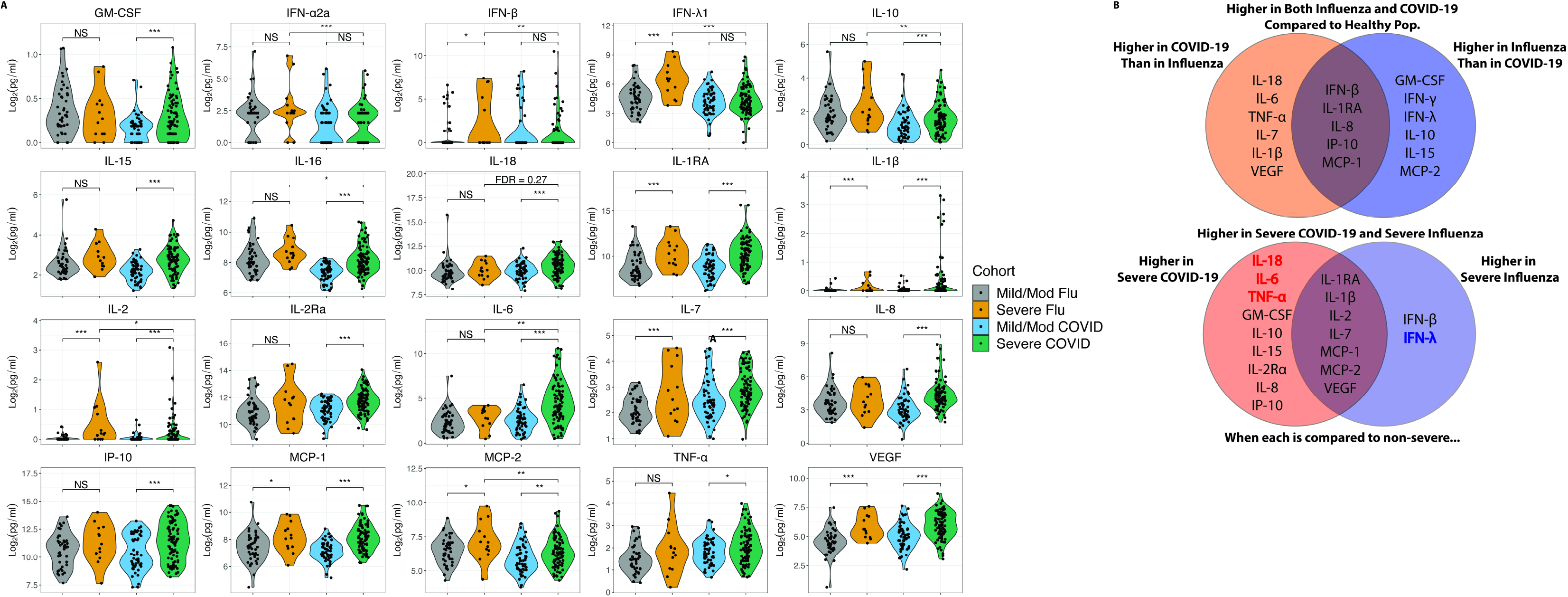
Cytokines and Chemokines Elevated in Severe Disease Compared to Mild/Moderate Disease and According to Infection. **A**. Differences between disease severity subgroups were determined by two-tailed t-test after adjusting for sex, age, and BMI. Differences between the severe COVID-19 cohort (green) and severe influenza cohort (orange) for each analyte were determined by two-tailed t-test after adjusting for sex, age, and BMI. False discovery rate (FDR) was obtained using the Benjamini-Hochberg procedure. Statistical significance is indicated by NS, *, **, or *** above the brackets indicating FDR >0.25, <0.25, <0.1, or <0.05 respectively. **B**. Top: Cytokines/chemokines higher in the COVID-19 cohort compared to influenza (left side), influenza compared to COVID-19 (right side) both COVID-19 and influenza compared to healthy controls, but not significantly different between COVID-19 and influenza (overlap center). Bottom: Cytokines/chemokines elevated in severe COVID-19 relative to mild/mod COVID-19 (left side), those elevated in severe influenza relative to mild/mod influenza (right side), and those that are elevated in severe forms of both diseases (overlap center).

When comparing severe COVID-19 to severe influenza directly, only IL-6 was significantly higher in severe COVID-19 whether adjusted for BMI or not (**Figure 3 and Supplementary Table 1 and 2**). IL-18 narrowly missed our pre-determined false discovery rate (FDR) cutoff for significance of 0.25 (FDR = 0.27) (**Supplemental Table 1 and 2**). Analytes higher in severe influenza compared to severe COVID-19 included IFN-λ1, IFN-α2a, IFN-β, IL-10, MCP-2, IL-16, and IL-2.

### IL-6 and IFN-λ1 Are the Most Important Cytokines in Distinguishing Severe COVID-19 from Severe Influenza

To further characterize potential differences in the inflammatory pathways activated by SARS-CoV-2 and influenza, we performed a multivariable analysis based on random forest using all the analytes and basic demographic information to compare severe COVID-19 and severe influenza. IL-6 and IFN-λ1 emerged as the most important factors distinguishing these two diseases in this analysis (**Figure 4**), with the highest fold changes between the severe groups. These findings underscore differences in the innate immune programs activated by these viruses; inflammatory macrophage activation pathways in COVID-19 and interferon pathways in influenza.

**Figure 4.**
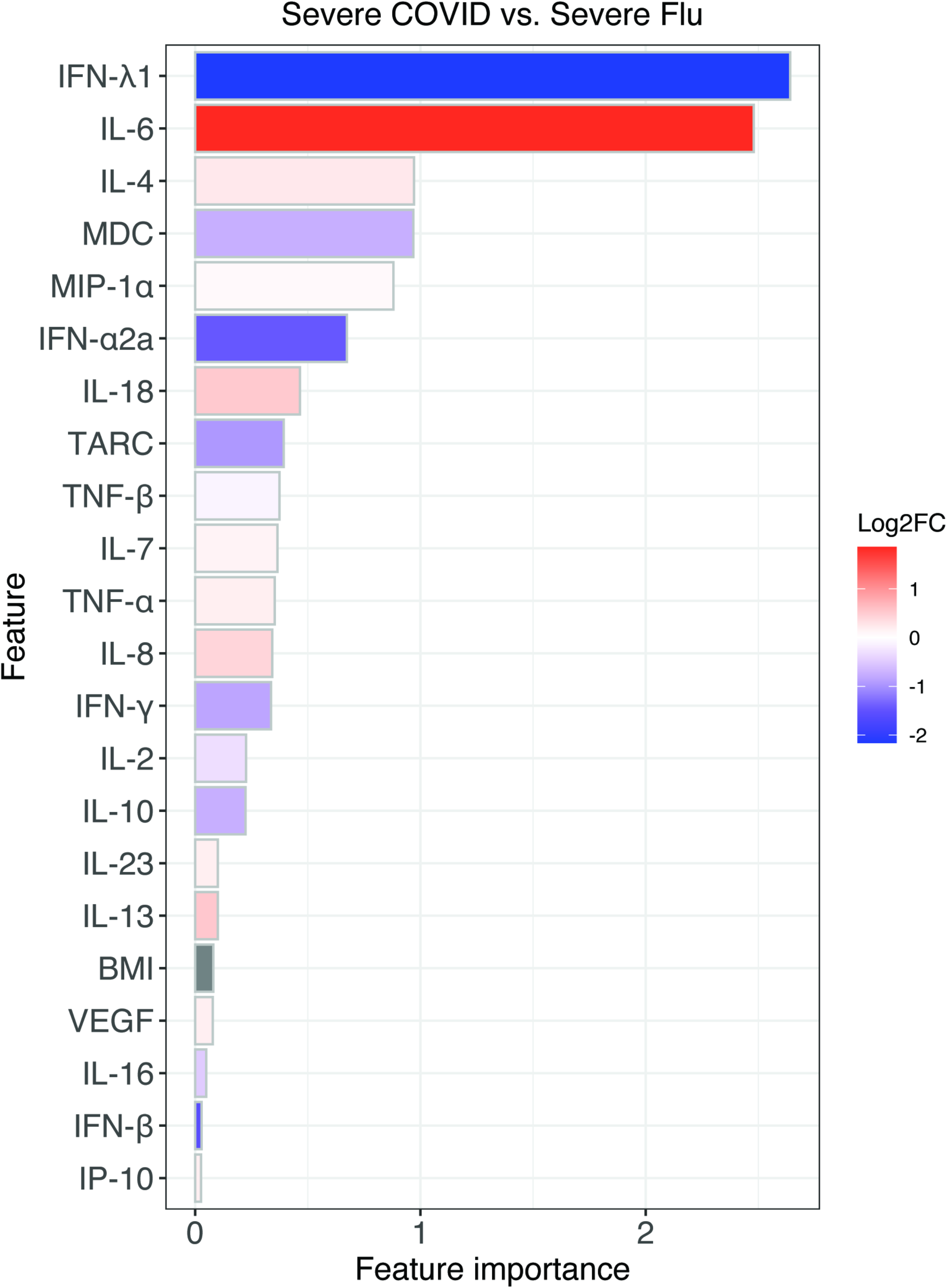
Multivariate analysis based on random forest revealed the most important variables in distinguishing severe COVID-19 and severe influenza. Feature importance was obtained from the random forest model for predicting severe COVID-19 vs. severe influenza. The color indicates the log_2_ fold change of the analyte signal between severe COVID-19 and severe influenza. Red color indicates a higher value in COVID-19 and blue color indicates a higher value in influenza.

### IL-1RA is a Potential Mediator of the Effect of BMI on COVID-19 Severity

Previous studies demonstrate an association between BMI and elevation of several of the cytokines we find increased in severe COVID-19, including IL-6, IL-1β, and IL-1RA [37,39,61,62]. Plotting BMI vs. cytokine concentration demonstrated a positive association between IL-1RA, IL-23p40, MDC, IL-17A, and MCP-2 (**Figure 5A and Supplemental Figure 4**). With mediation analysis and after adjusting for multiple testing, only IL-1RA had a significant mediation effect. IL-1RA is an acute phase reactant produced by macrophages and the liver in response to inflammatory cytokines and pathogens through pathways that upregulate IL-6 and TNF-α [63,64] While the total effect of BMI on COVID-19 severity was 0.008 (FDR < 0.1), most of the effect was indirectly through IL-1RA. Indeed, the direct effect of BMI on COVID-19 severity was minimal (−0.0003, FDR=0.93) compared to the indirect effect of BMI on severity through IL-1RA (0.0083, FDR < 0.05). This suggests that the effect of increased BMI on the likelihood of severe COVID-19 may be mediated by IL-1RA levels (**Figure 5B, Supplementary Table 3**). A similar analysis of the influenza cohort did not reveal any cytokines/chemokines mediating BMI and disease severity (**Supplementary Table 4**)..

**Figure 5.**
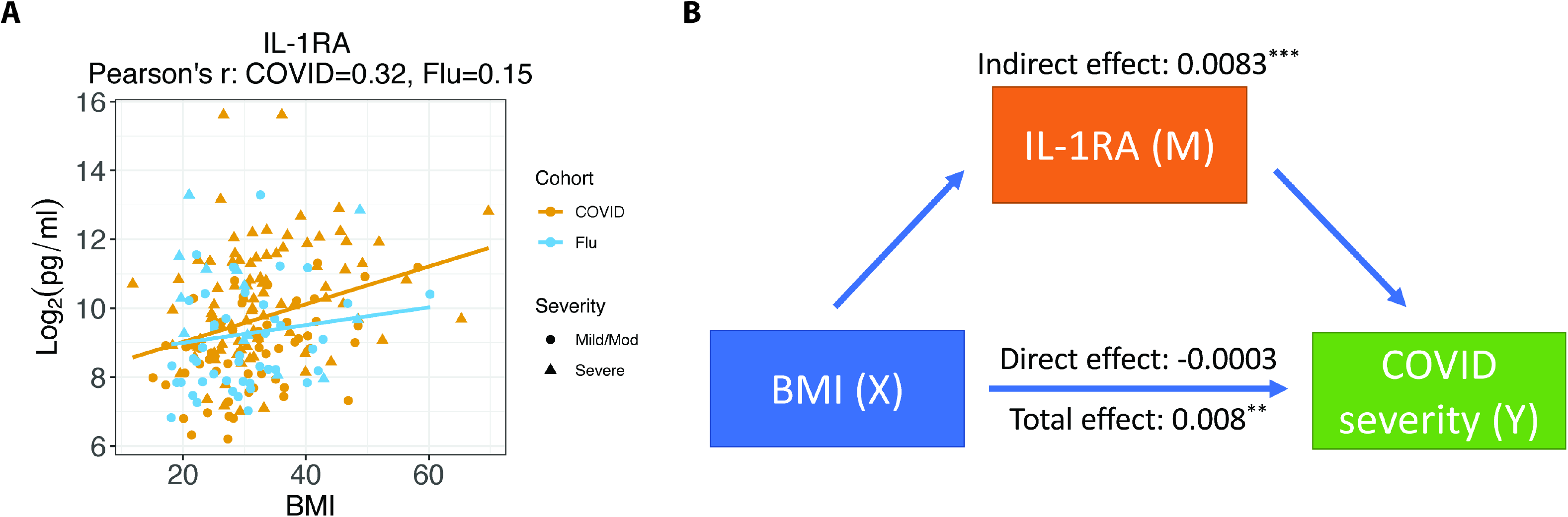
Mediation Analysis of BMI and IL-1RA on COVID-19 Severity. **A**. BMI (X axis) was plotted vs. IL-1RA level (Y axis). The yellow line is a linear regression line for the COVID-19 cohort and the blue line is a linear regression line for the influenza cohort. The Pearson’s correlation coefficient between IL-1RA and BMI is 0.32 for the COVID-19 cohort and 0.15 for the influenza cohort. **B**. Diagram of the mediation analysis of BMI and IL-1RA on COVID-19 severity. The indirect, direct, and total effects were showed in the diagram. Statistical significance is indicated by *, **, or *** representing FDR<0.25, <0.1, or <0.05 respectively.

## Discussion

Our analysis of cytokines and chemokines elevated in COVID-19 compared to influenza reveals distinct cytokine profiles of these respiratory diseases. We found that several cytokines previously reported to be elevated in COVID-19 were either not different from influenza or actually more elevated in influenza than in COVID-19 including IFN-γ, IL-10, and IL-15 [15]. While some have proposed that targeting these cytokines may be beneficial in COVID-19, our data demonstrating comparatively higher levels in moderate severity influenza do not support this [58].

Another study of cytokine profiles in COVID-19 and influenza also found that many cytokines were either not different or were significantly less elevated in COVID-19 versus influenza [65]. Consistent with their results, we found that IFN-γ was elevated in influenza. They found significant increases in IL-1RA, IL-2, IL-17, and MIP-1α in their influenza cohort relative to COVID-19, while we did not. These differences may be due to our adjustment for BMI, our larger cohort size, different platforms used to measure cytokines, or the fact that a higher percentage of their influenza patients had severe disease. That same study also found a trend toward increased IL-6 and IL-8 in the COVID-19 group compared to influenza and we found that both were significantly increased when comparing COVID-19 to influenza. They did not measure IL-18 or IFN-λ1, which were important in distinguishing severe COVID-19 from severe influenza in our analysis. Importantly, both this study and ours failed to identify a non-specific “cytokine storm” in COVID-19 subjects. While the “cytokine storm” hypothesis was put forth at the beginning of the pandemic [66] to explain the pathology observed in COVID-19, our study demonstrates lower overall cytokine levels in COVID-19 than those observed in other inflammatory diseases [28,29].

Despite the fact that both influenza and SARS-CoV-2 are respiratory RNA viruses, our study emphasizes that they induce distinct inflammatory pathways. We found that severe COVID-19 leads to upregulation of cytokines associated with a proinflammatory macrophage phenotype [67] characterized by high levels of IL-6, TNF-α, and IL-18, whereas interferons and cytokines involved in T cell activation (IL-15 and IFN-γ) are upregulated in influenza. IL-18 and IL-1β (which is difficult to detect in blood) are released upon activation of a component of the innate immune system called the inflammasome, often activated in macrophages [53,55,68]. Emerging evidence suggests inflammasome activation is central to the pathogenesis of SARS-CoV-2 and is a marker of severe disease [8,12,69,70]. Our study provides additional evidence that this pathway is activated in SARS-CoV-2 infection. Whether this is simply a marker of severity or contributes to the COVID-19 pathology remains to be determined.

High IL-6 and low IFN-λ1 were the most distinct features of severe COVID-19 compared to severe influenza, consistent with results of another study of COVID-19 and influenza patients [57]. It is important to note that we do not know if these cytokine disparities mediate the pathology differences, or rather are merely correlates of other distinct immune responses. The immune response to SARS-CoV-2 is complex and while steroids have proven beneficial in later stages of the disease in patients requiring oxygen, immunosuppression early is not beneficial [71]. Given the association with severe COVID-19 and a proinflammatory macrophage phenotype, targeting of the cytokines mediating MAS, singly or in combination, might provide a more specific approach to immune modulation. Indeed, depending perhaps on timing of administration, targeted anti-IL-6 therapy has been associated with clinical benefit[25,26,72].

Another strength of our analysis is the focus on BMI as a contributor to severe disease. While this association has been widely described, few analyses of inflammatory cytokines have taken this variable into account [73]. We found that IL-1RA is a potential mediator of the effect of BMI on COVID-19 disease severity. This novel observation points to a possible mechanism linking BMI to severe COVID-19. Adipocytes are a major source of IL-1RA [39] and elevated BMI provides more adipose tissue to produce this acute phase reactant. This was an unexpected finding given the anti-inflammatory nature of this cytokine. The role of IL-1RA in pathogenesis warrants additional investigation, particularly in light of studies reporting benefit with the IL-1RA analog, anakinra, in COVID-19 [17,18].

Limitations to our study include that outpatients with mild COVID-19 were not included. However, we would predict that differences between those with and without severe disease in our study would only be more significant if milder disease were included. In addition, our study subjects with influenza also had disease severe enough to warrant hospitalization, making them an appropriate comparator. An additional limitation is that we examined a single timepoint. It is possible that the dynamics of these cytokines during the course of hospitalization would change in such a way that the cytokine profiles would become even more or less distinct from influenza. However, as patients remain hospitalized, complications from critical illness arise that could obfuscate the comparison to influenza. Finally, the influenza cohort was, on average, younger than the COVID-19 cohort. However, we adjusted for age in our analysis.

This study provides insight into pathways activated by SARS-CoV-2 and influenza, demonstrating that some inflammatory cytokines elevated in COVID-19 likely reflect common pathways activated in respiratory tract inflammation, while others are more specific to COVID-19 pathogenesis. We identified IL-1RA as a potential mediator of the effect of increased BMI on COVID-19 disease severity. In summary, this study demonstrates activation of a proinflammatory cytokine macrophage pathway and a role for IL-1RA in severe COVID-19, highlighting potential therapeutic targets.

## Data Availability

Anonymized data will be available upon reasonable request to the corresponding author.

## ACKNOWLEDGMENTS

We would like to acknowledge the contribution of the Johns Hopkins IVAR team and assistance for clinical data coordination and retrieval from the Core for Clinical Research Data Acquisition. The specimens from COVID-19 patients utilized for this study were part of the Johns Hopkins Biospecimen Repository, which is based on the contribution of many patients, their families, research teams, and clinicians. We thank members of the Viral Hepatitis Center at Johns Hopkins for advice and discussion and blood processing. We owe additional thanks to Sherry Kelly Gibson, Muhammad Munir, Tingtin Niu, Nikitta Dhillon, Dan Warren, Robin Avery and Niraj Desai for assistance with the study.

## FUNDING

This work was supported by the Johns Hopkins COVID-19 Research Response Program, a Johns Hopkins University Provost Research Grant, The Bill and Melinda Gates Foundation (134582), National Cancer Institute (U54CA260491), and National Institutes of Health Centers of Excellence in Influenza Research and Surveillance (HHSN272201400007C). The study was supported in part by a cooperative agreement between Johns Hopkins University (JHU) and the Division of Intramural Research, National Institute of Allergy and Infectious Diseases, the US Department of Health and Human Services Biomedical Advanced Research and Development Authority (BARDA; agreement number IDSEP160031-01-00), the National Institute of Allergy and Infectious Diseases U19AI088791 and R01AI108403 and National Institute of Allergy and Infectious Diseases contract HHSN272201400007C awarded to Johns Hopkins Center of Excellence in Influenza Research and Surveillance (JHCEIRS). HJ and WZ were supported in part by the National Institute of Health grant R01HG009518. AHK was supported by the National Institute of Health T32 AI007291-27. AF was supported, in part, by grant D18HP29037 from the U.S. Health Resources and Services Administration, Bureau of Health Workforce, Health Careers Opportunity Program. RER was support in part by the National Institute of Allergy and Infectious Diseases contract HHSN272201400007C awarded to the Johns Hopkins Center of Excellence in Influenza Research and Surveillance (JHCEIRS) at the Johns Hopkins University and US Department of Health and Human Services Biomedical Advanced Research and Development Authority (BARDA; agreement number IDSEP160031-01-00).

## Conflicts of Interest

None of the authors have any relevant conflicts of interests to disclose.

## Supplementary Methods

*P*-values for comparisons of age and BMI between COVID-19 and influenza cohorts were determined by a Wilcoxon-Rank-Sum test. *P*-values for differences in sex and non-white race between COVID-19 and influenza were determined by a Fisher’s exact test. Ethnicity data was not collected in the influenza study and therefore race was treated as a binary variable (white or not white) for purposes of this comparison.

The cytokine/chemokine (i.e., analyte) signals were first log2 transformed after adding a pseudocount of one. To compare the analytes between patient groups, a linear regression model was fitted for each analyte to test the difference between every pair of patient groups (e.g., COVID-19 vs. influenza) after adjusting for covariates (e.g., age, sex, and BMI). For instance, in the model comparing COVID-19 and influenza patients, the analyte signal was treated as the dependent variable and the patient group (i.e., COVID-19 or Influenza), age, sex, and BMI were treated as independent variables. P-values of the coefficient for the patient group from the model were obtained and converted to false discovery rates (FDR) using the Benjamini-Hochberg (BH) procedure. **Figure 1** and **Supplementary Figure 1** show the comparisons between COVID-19 and healthy controls, influenza and healthy controls, and COVID-19 and influenza. **Figure 3** and S**upplementary Figure 3** show the comparisons between severe and mild/mod COVID-19, severe and mild/mod influenza, and severe COVID-19 and severe influenza.

To further examine cytokines/chemokines that drive the difference between the two diseases, a multivariate analysis using R packages caret [45] and randomForest [46] based on a machine learning model, random forest were utilized. First a random forest model was trained using the analyte signals, age, sex, and BMI as features based on leave-one-sample-out cross-validation for predicting severe COVID-19 vs. severe influenza patients. Then, the feature importance was calculated based on the mean decrease of accuracy that measures how much a feature affects the prediction accuracy. Since the number of severe influenza patients (N = 13) is smaller than the number of severe COVID-19 patients (N = 89), 13 severe COVID-19 patients were selected at random to match the sample size between the two groups. This was repeated ten times and the average of feature importance from the ten experiments was reported.

The connection among BMI, cytokine/chemokine, and disease severity was studied by applying a mediation analysis using R package mediation [47]. Each analyte was assessed as a potential mediator in the pathway between BMI and COVID-19 severity. The significance of the indirect effect was tested using the nonparametric bootstrap method in the package. The indirect effects, direct effects, total effects and the associated p-values, confidence intervals were calculated. To adjust for multiple testing, p-values for the indirect effects and direct effects were converted to FDR using the BH procedure. A similar analysis was applied to the influenza cohort.

---

**Supplemental Figure 1. Cytokines and Chemokines Elevated in Influenza and COVID-19 Compared to Healthy Controls (HC)**.

Differences between HC and the COVID-19 cohort or influenza cohort were determined by two-tailed t-test after adjusting for sex and age. Differences between the COVID-19 cohort and influenza cohort for each analyte were determined by two-tailed t-test after adjusting for sex, age, and BMI. FDR was obtained using the Benjamini-Hochberg procedure. Statistical significance is indicated by NS, *, **, or *** above the brackets indicating FDR >0.25, <0.25, <0.1, or <0.05 respectively.

**Supplemental Figure 2. Cytokine and Chemokine Correlation Matrix using both COVID-19 and influenza patients**.

Correlations between each analyte and every other analyte were determined by Pearson’s correlation coefficient of each pair of cytokine/chemokine across all COVID-19 and influenza patients. The heat bar on the right side of the figure indicates both the directionality of the correlation (blue represents a positive correlation and red represents a negative correlation) and the strength (darker values indicate a correlation coefficient closer to 1 or −1).

**Supplemental Figure 3. Cytokines and Chemokines Elevated in Severe Disease Compared to Mild/Moderate Disease**.

Differences between disease severity subgroups were determined by two-tailed t-test after adjusting for sex, age, and BMI. Differences between the severe COVID-19 cohort and severe influenza cohort for each analyte were determined by two-tailed t-test after adjusting for sex, age, and BMI. FDR was obtained using the Benjamini-Hochberg procedure. Statistical significance is indicated by NS, *, **, or *** above the brackets indicating FDR >0.25, <0.25, <0.1, or <0.05 respectively.

**Supplemental Figure 4. Correlation between cytokines/chemokines with BMI in COVID-19 and Flu**.

The scatter plots show the correlation between cytokines/chemokines with BMI in COVID-19 (yellow dots) or Flu (blue dots) patients. Linear regression lines were fitted between the signals of an analyte and BMI for COVID-19 and Flu patients, respectively. Pearson’s correlation coefficient between an analyte and BMI for COVID-19 or Flu patients is indicated in the plot.

**Supplemental Table 1. Coefficients, p values and FDR for all comparisons without BMI adjustment**.

**Supplemental Table 2. Coefficients, p values and FDR for all comparisons with BMI adjustment**.

**Supplemental Table 3. Mediation Analysis of BMI and cytokines/chemokines on disease severity for COVID-19**.

**Supplemental Table 4. Mediation Analysis of BMI and cytokines/chemokines on disease severity for influenza**.

## Notes

### Competing Interest Statement

The authors have declared no competing interest.

### Author Declarations

All studies were approved by the Johns Hopkins (JH) Institutional Review Board. Hospitalized patients diagnosed with COVID-19 by positive SARS-CoV-2 RNA testing in the Johns Hopkins Healthcare System were enrolled in a prospective consented protocol to investigate research questions specific to the clinical course of COVID-19 (JH IRB 00245545). Plasma samples from hospitalized patients infected with influenza between 2017 and 2019 were obtained as previously described for comparison to hospitalized COVID-19 patients in this study [41,42], under an IRB approved protocol (JH IRB 00091667). The healthy control (HC) plasma samples were obtained from HIV/HCV-antibody seronegative subjects enrolled before 2020 in the Baltimore Before and After Acute Study of Hepatitis (BBAASH) study (JH IRB NA_00046368)

