## Supplementary figures and images for "Differential Cytokine Signatures of SARS-CoV-2 and Influenza Infection Highlight Key Differences in Pathobiology"

### SupplementalFig2.tif

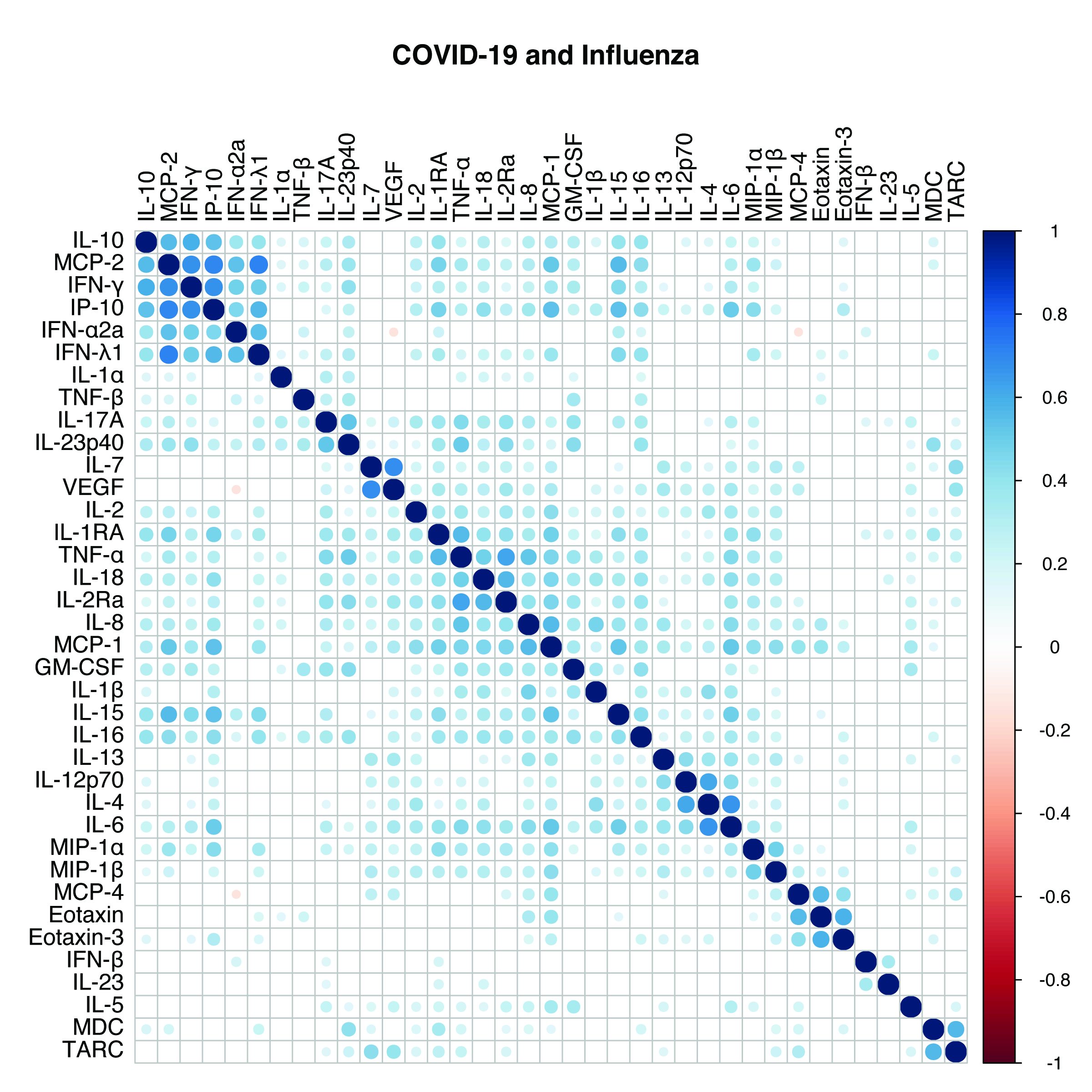
